# Analytical Performance of the AMDI™ Fast PCR Mini Respiratory Panel

**DOI:** 10.64898/2025.12.22.25342853

**Authors:** Louis Geller, Alyssa Fiore, Liza Perez, Corinne Minzer, Yujia Liu, Estibaliz Alvarado, Nicholas Casarez, Brenda Martinez, Justin Nako, Gabriel Rodriguez, Yvette Arroyo, Nicholas Johnsen, Christopher Ross, Francesca Foltz, Regina Martin, Regis Peytavi, Aravind Srinivasan

**Affiliations:** Autonomous Medical Devices Incorporated, Santa Ana, CA

## Abstract

Influenza A, Influenza B, SARS-CoV-2, Respiratory Syncytial Virus and other respiratory pathogens are an ongoing public health concern. The ability to rapidly identify these viruses early in infection is essential for effective treatment and outbreak control. The AMDI™ Fast PCR Mini Respiratory Panel (MRP) incorporates sample preparation and real-time RT-PCR for detection of Flu A, Flu B, SARS-CoV-2 and RSV from anterior nasal swab (ANS) specimens in less than 10 minutes at the point of care. We established the analytical performance characteristics of the Fast PCR MRP and determined that the limit of detection (LoD) is 250 copies/mL for Flu A and RSV, and 500 copies/mL for Flu B and SARS-CoV-2. In a reproducibility study at 3 clinical sites, there was at least 98.2% positivity for each target for a weak positive (2x LoD) sample, at least 99.6% positivity for a moderate positive (5x LoD) sample and the negative sample returned a negative result for at least 99.3% of the tests. Fast PCR MRP had 100% analytical reactivity to all strains tested (23 Flu A, 6 Flu B, 8 SARS-CoV-2 and 6 RSV) and at least 98% predicted inclusivity from *in silico* analysis. There was no cross-reactivity to 40 viruses, bacteria and fungi, nor interference for 15 endogenous and exogenous substances in ANS matrix. The Fast PCR MRP delivers excellent analytical performance comparable to high complexity laboratory assays, at the point of care.

## Introduction

Respiratory viruses such as Influenza, RSV and COVID-19 are a major cause of severe illness and socioeconomic burden especially among vulnerable populations (1, 2). Laboratory methods for the detection of respiratory viruses have evolved and improved considerably over the past 10 years with respect to sensitivity, specificity and turnaround time (3). Previously, cell culture and immunofluorescence-based methods, such as direct fluorescent antibody (DFA), were widely used for antigen detection. Rapid antigen detection assays, including lateral flow methods, are used today for point of care (POC) tests; however, while these offer fast turnaround time, they often require interpretation by a visual readout and lack the required sensitivity leading to a high rate of false negatives (3–4). Due to increased sensitivity, multiplex capability, and the ability to detect viral nucleic acids earlier than antigens, Nucleic Acid Amplification Tests (NAATs) including PCR-based assays are now the gold standard for diagnosis of respiratory viral infections, such as Influenza A (Flu A), Influenza B (Flu B), Respiratory Syncytial Virus (RSV) and COVID-19 (SARS-CoV-2) (3, 5–7). While rapid antigen tests and NAATs both have their merits, the public health crisis during the global COVID-19 pandemic underscored their drawbacks and highlighted an unmet need for accessible and simple molecular tests that provide results at the speed of less-sensitive antigen tests with the same sensitivity as high-complexity NAATs.

The AMDI™ Fast PCR Mini Respiratory Panel (MRP) is a multiplex real-time reverse transcriptase PCR (RT-PCR) test for Flu A, Flu B, RSV and SARS-CoV-2 from human anterior nasal swab specimens and is designed as a CLIA-waivable POC test. The test automates sample to answer in less than 10 minutes, providing a faster option than rapid antigen tests while preserving the quality of PCR assays. The Fast PCR system employs a novel sample processing method using hyperbaric heating to lyse microorganisms and neutralize PCR inhibitors within seconds, followed by amplification using RT-PCR, and cloud-connected data analysis to generate the result. We recently published the results of a multicenter clinical study for the Fast PCR MRP test (8). This study, conducted at 9 clinical sites, included 1906 patient samples and compared the performance of the Fast PCR MRP to the Cepheid Xpert® Xpress CoV-2/Flu/RSV plus. This study demonstrated excellent clinical performance of the Fast PCR MRP test.

FDA-registered medical devices incorporating RT-PCR assays for detecting respiratory viruses, including the Fast PCR MRP, usually have very good clinical sensitivity and specificity established through clinical agreement studies. By design, prospective clinical studies provide a snapshot of the ability of a test to detect circulating strains in specific geographic regions. Clinical validation studies are performed in the intended test environment; however, these studies use sample cohorts for which there is often no information about viral loads or subtypes/strains that are prevalent at the time of testing. To augment such prospective clinical studies, analytical studies for POC tests are typically executed at the manufacturer’s facility by trained operators, with known strains and concentrations. These studies provide complementary information including analytical sensitivity, reproducibility, analytical specificity, and interference. Data from such controlled studies could be valuable in determining if a given test addresses the specific testing needs of a laboratory, hospital, or clinic. In addition, this information is useful for sensitivity comparisons with other similar systems.

In this paper, we present and discuss the analytical performance characteristics including limit of detection, reproducibility, inclusivity and cross-reactivity of the Fast PCR MRP test. In addition, we also evaluated the potential for interference from microorganisms, endogenous substances, and common medications used to alleviate symptoms from respiratory infections.

## Materials and Methods

### Sample Matrix

Anterior Nasal Swab (ANS) specimens were collected from volunteers under an institutional review board-approved protocol (Pearl IRB Study ID# 21-AMDI-101), eluted in AMDI Sample Buffer (Santa Ana, CA; Part # AMDI-0005), pooled and screened to confirm negative status for the viral targets in the Fast PCR MRP assay.

### Microorganism Stocks

Microorganism stocks were obtained from Microbiologics (San Diego, CA), ZeptoMetrix (Buffalo, NY) and ATCC (Manassas, VA). Viral, bacterial and fungal stocks were quantified by droplet digital PCR (ddPCR) and/or titered in TCID_50_/mL, CFU/mL or CCU/mL by the vendor.

### Contriving of Samples

Test samples were contrived by serial dilution of the microorganism stocks in negative ANS matrix to the appropriate concentration, while keeping organism stocks, negative matrix and samples on ice. Contrived samples were stored below −70 °C until further testing.

### Testing of Samples

All samples were tested using the AMDI Fast PCR MRP kit on the Fast PCR system (Figure 1) that includes the Fast PCR Operating Module and Fast PCR Base Station from AMDI, using the quick reference guide. Samples were thawed at room temperature and placed at 2-8 °C. Samples were warmed to room temperature for at least 10 minutes immediately prior to testing. For testing, 400 µL of the sample was loaded into the sample port of the Fast PCR MRP Test Disc using a transfer pipette, and the sample port cover was closed. The operator scanned the Test Disc ID barcode using the Fast PCR Operating Module and the disc was loaded into the instrument to start the test. The Fast PCR Operating Module and test disc processed the sample by hyperbaric heating, followed by microfluidic metering of the processed sample, and real-time RT-PCR. Data analysis of raw fluorescence signal and automatic result generation is completed in the AMDI Cloud and displayed on the tablet connected to the Fast PCR System.

**Figure 1:**
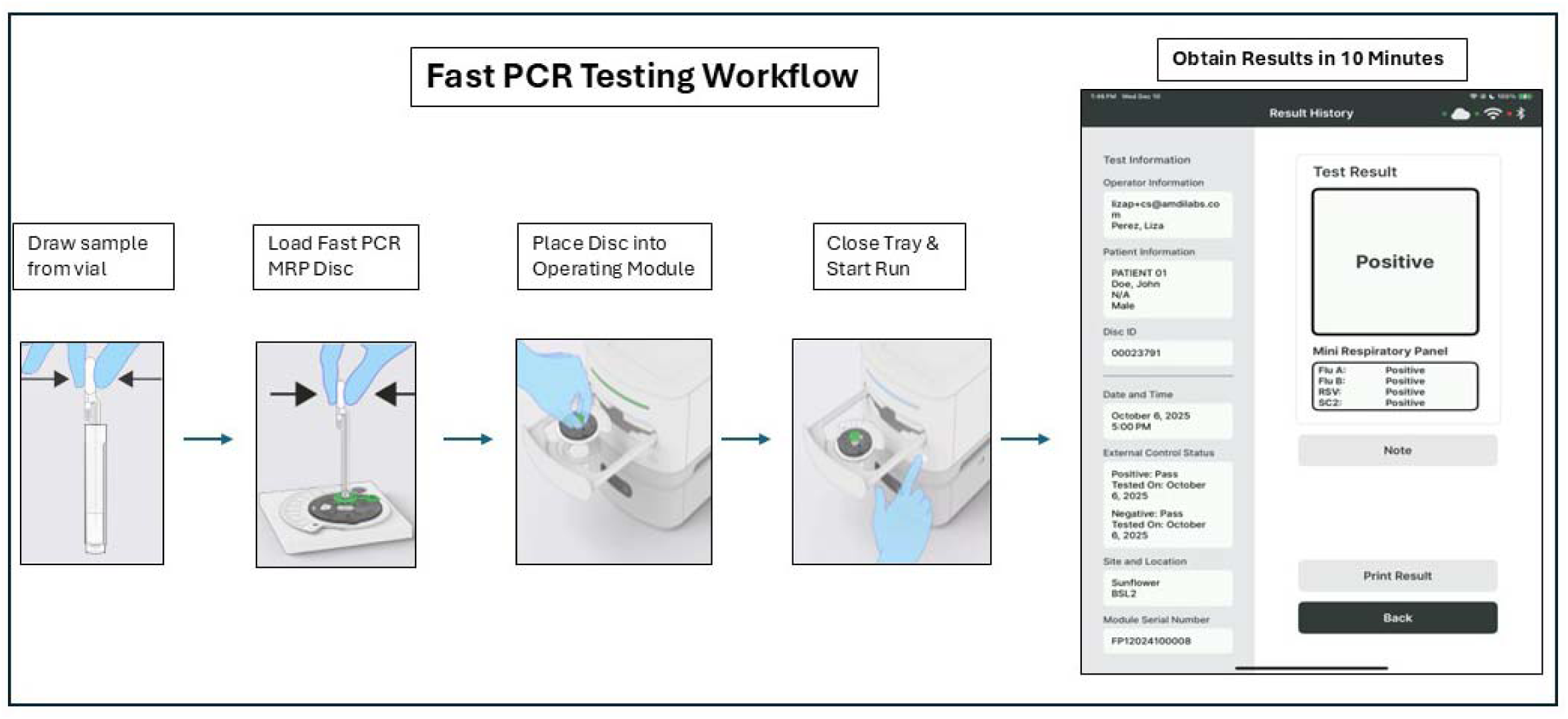
Testing of a sample with the Fast PCR MRP kit includes drawing the sample from the vial using a transfer pipette, loading a Fast PCR MRP Disc, placing the Disc into the Fast PCR Operating Module and closing the tray to start the run. The Fast PCR System performs sample preparation by hyperbaric heating, real-time RT-PCR and analysis. In approximately 10 minutes, the results are uploaded to the AMDI Cloud and displayed on the iPad.

### Quality Control

Quality control materials for the Fast PCR System consisted of lyophilized positive and negative external controls, which were reconstituted with AMDI Sample Buffer and run on every Fast PCR Instrument on each day of testing.

### Limit of Detection (LoD) Determination

The viral strains used for the LoD determination are shown in Table 1. The preliminary LoD (lowest level with 100% positivity) was determined by testing each viral strain separately in negative matrix at 2250, 750, 250, 83 and 27 copies/mL, in triplicate, using one lot of Fast PCR MRP kit. For confirmation, 20 replicates of the viral strain at its preliminary LoD were prepared and tested, along with 20 replicates each of contrived samples above and below the preliminary LoD, using two lots of Fast PCR MRP kits. The LoD was defined as the lowest level with at least 95% positivity.

**Table 1:**
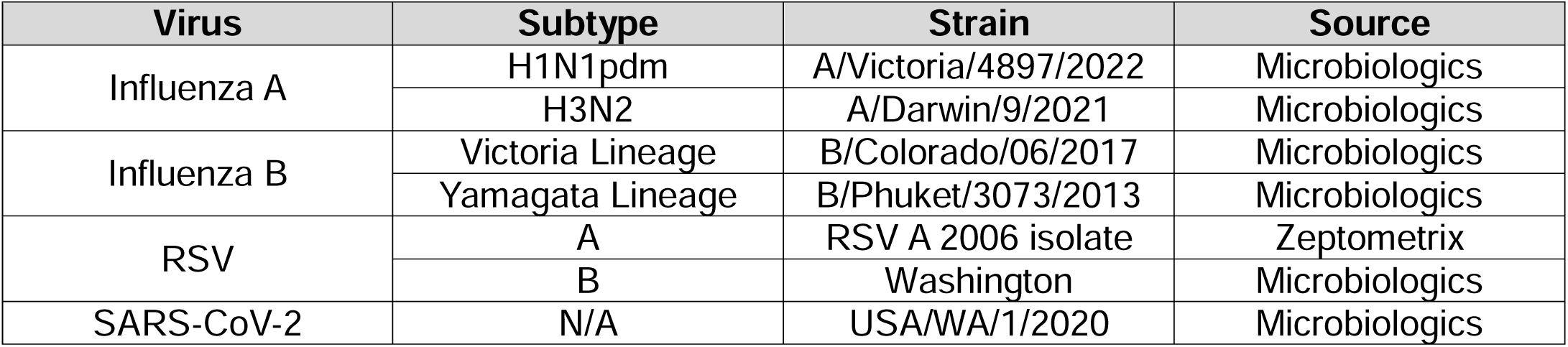
Viral strains used for determining the LoD of each viral target in the Fast PCR MRP Test.

Once the LoD was determined for each viral target, a quadspike sample containing 1 strain of each virus at its LoD was prepared, and 20 replicates were tested. The quadspike LoD sample was composed of the Influenza A/Darwin/9/2021, Influenza B/Colorado/06/2017, RSV A 2006 isolate and SARS-CoV-2/USA/WA1/2020 strains.

### Multi-site Reproducibility

A reproducibility study was conducted at three CLIA-waived testing sites, with two untrained operators per site. Each site evaluated three independent lots of the Fast PCR MRP kit and three Fast PCR Operating Modules/Base Station units, over the course of at least five non-consecutive days. The sample panel consisted of three samples in ANS matrix: weak positive (2x LoD) quadspike, moderately positive (5x LoD) quadspike, and negative matrix. The 2x and 5x LoD quadspike samples were contrived using Influenza A/Darwin/9/2021, Influenza B/Colorado/06/2017, RSV A 2006 isolate and SARS-CoV-2/USA/WA1/2020. At each clinical site, at least 90 replicates per sample were tested. In addition, three different lots of the Fast PCR MRP Positive External and Negative External Controls were evaluated by each of the sites. These controls were tested on each instrument prior to testing any samples in the reproducibility panel.

### Inclusivity (Analytical Reactivity)

Table S4 lists the viral strains tested for analytical reactivity for each target in the Fast PCR MRP Test. Each viral strain was diluted to 3x LoD of the respective subtype in negative ANS matrix and tested in triplicate. For Flu A, Flu B and RSV, strains were diluted to 3x LoD, according to the subtype of the strain to be tested. For SC2, dilutions were prepared based upon the LoD for the USA/WA1/2020 strain. For viral stocks quantified using ddPCR, the stock was diluted to 3x LoD based upon copies/mL; otherwise, stocks were diluted to 3x LoD as measured in TCID_50_/mL.

In addition to laboratory testing of viral strains for inclusivity, *in silico* analysis for analytical reactivity was also performed. For SARS-CoV-2, this included sequence retrieval and alignment from GISAID (Global Initiative on Sharing All Influenza Data [9]) for sequences prior to 06-11-2024, and NCBI Virus (10) followed by Geneious Prime (Dotmatics, Boston, MA) for sequences newer than 06-11-2024. For Flu A, Flu B and RSV, NCBI Virus and Geneious Prime were used to retrieve viral sequences covering the entire target region, from a human host. Following alignments, a Python script was used for the analysis. For each viral target, no more than 2 mismatches were allowed per oligonucleotide, as well as no mismatches in the three 3’ nucleotides of each primer.

### Cross-reactivity

Table S5 lists the microorganisms evaluated for cross-reactivity in the Fast PCR MRP Test. For this evaluation, each organism was diluted in negative ANS matrix either at 1×10^6^ CFU/mL or CCU/mL (for bacteria or fungi) or 1×10^5^ TCID_50_/mL, PFU/mL or copies/mL (for viruses) and tested in triplicate. For organisms where stocks of sufficient titer could not be obtained, testing was performed at the highest possible concentration.

In addition to laboratory testing of organisms for cross-reactivity, *in silico* analysis/organism-specific BLAST (11) was also performed using the oligonucleotides for each target as the QUERY. The blastn program, Core nucleotide database (core_nt) and default parameters were selected, allowing BLAST to automatically adjust parameters for short input sequences. For each target, the intended organism was excluded from the analysis. BLAST results were downloaded in XML format and analyzed using a Python script. For an assay to be considered cross-reactive to an off-panel organism, each oligonucleotide of a given assay was required to have at least 80% homology to the genome of that organism. Primers were required to have less than 4 mismatches in the 3’ end, anneal to the genome of that organism within 200 base pairs of each other and have the ability to extend. Further, all oligonucleotides for a given assay must anneal to the genome of the cross-reacting organism in the proper orientation, with the probe annealing between the primers, to be considered cross-reactive.

### Interference

Potentially interfering substances (Table S6) were dissolved in AMDI Sample Buffer and used to prepare working stocks. A 3x LoD quadspike was prepared in negative ANS matrix and mixed with each substance under evaluation, at the test concentration. For microbial interference, a 3x LoD quadspike was prepared in negative ANS matrix and mixed with each organism being evaluated (Table S5) at the same concentration used for cross-reactivity (1×10^6^ CFU/mL or CCU/mL for bacteria or fungi and 1×10^5^ TCID_50_/mL, PFU/mL or copies/mL for viruses).

Samples containing potentially interfering substances or off-panel organisms were tested in triplicate. If less than 100% positivity was observed, the concentration of the substance or organism was lowered and retested, until there was 100% positivity.

### Data Analysis

Calculations were performed using Microsoft Excel, GraphPad Prism 10 (GraphPad Software, Boston, MA) and JMP18 (JMP Statistical Discovery LLC, Cary NC). A custom Python script was used for analysis of *in silico* analytical reactivity and cross-reactivity data.

## Results

### Limit of Detection (LoD)

The LoD was determined for two Influenza A strains, two Influenza B strains, two RSV strains and one SARS-CoV-2 strain. Strain specific LoDs, Ct values at LoD are summarized in Table 2 and Table S1.

**Table 2:**
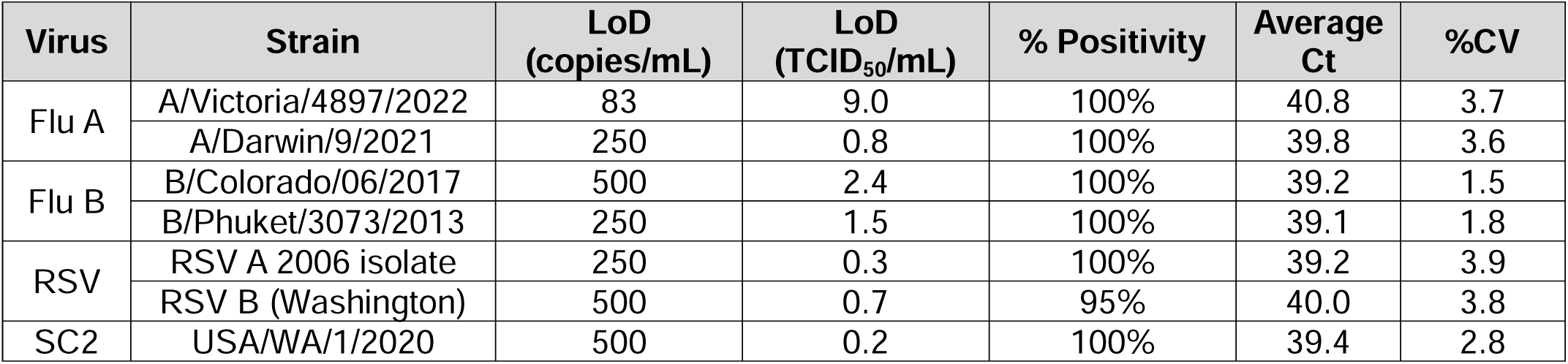
Limit of Detection Results.

A quadspike sample with one strain of each virus at 1x LoD showed 100% positivity, and no significant difference in Ct values (Table 3).

**Table 3:** Quadspike LoD Results and Comparison to Singlespike LoD.

| Virus | Strain | copies/mL | % Positivity | Average. Ct |  |  |
| --- | --- | --- | --- | --- | --- | --- |
|  |  |  |  | Singlespike | Quadspike | p-value |
| Flu A | A/Darwin/9/2021 | 250 | 100% | 39.8 | 39.6 | > 0.05 |
| Flu B | B/Colorado/06/2017 | 500 | 100% | 39.2 | 38.9 | > 0.05 |
| RSV | RSV A 2006 isolate | 250 | 100% | 39.2 | 39.2 | > 0.05 |
| SC2 | USA/WA/1/2020 | 500 | 100% | 39.4 | 38.9 | > 0.05 |

### Multi-site Reproducibility

A Reproducibility study was conducted across three CLIA-waived clinical sites, with three samples: weak positive (2x LoD), moderately positive (5x LoD) and negative. For the 2x LoD sample, there was at least 98.2% agreement between expected and observed results for each viral target; for the 5x LoD sample, there was at least 99.6% agreement; and for the negative sample, there was at least 99.3% agreement (Table 4 and Table S2).

**Table 4:** Percent Agreement for Multi-Site Reproducibility.

| Sample | Analyte | Negative | Positive | Total | Agreement | Lower 95% CI | Upper 95% CI |
| --- | --- | --- | --- | --- | --- | --- | --- |
| 2x LoD | Flu A | 3 | 270 | 273 | 98.9% | 96.8% | 99.6% |
|  | Flu B | 1 | 272 | 273 | 99.6% | 98.0% | 99.9% |
|  | RSV | 5 | 268 | 273 | 98.2% | 95.8% | 99.2% |
|  | SC2 | 3 | 270 | 273 | 98.9% | 96.8% | 99.6% |
| 5x LoD | Flu A | 0 | 272 | 272 | 100.0% | 98.6% | 100.0% |
|  | Flu B | 0 | 272 | 272 | 100.0% | 98.6% | 100.0% |
|  | RSV | 1 | 271 | 272 | 99.6% | 97.9% | 99.9% |
|  | SC2 | 0 | 272 | 272 | 100.0% | 98.6% | 100.0% |
| Negative | Flu A | 276 | 2 | 278 | 99.3% | 97.4% | 99.8% |
|  | Flu B | 278 | 0 | 278 | 100.0% | 98.6% | 100.0% |
|  | RSV | 278 | 0 | 278 | 100.0% | 98.6% | 100.0% |
|  | SC2 | 277 | 1 | 278 | 99.6% | 98.0% | 99.9% |

In addition, there was 100% agreement between expected and observed results for daily QC at all sites, using the External Positive and External Negative Controls (Table S3).

### Inclusivity (Analytical Reactivity)

Inclusivity was evaluated by testing 23 Influenza A strains (7 H1N1pdm, 5 seasonal H1N1, 11 H3N2), 6 Influenza B strains (4 Victoria Lineage and 2 Yamagata Lineage), 6 RSV strains (3 RSV A and 3 RSV B), and 8 SARS-CoV-2 strains, in addition to the seven strains tested during the LoD determination. All strains had 100% positivity (3/3 replicates) at 3x LoD (Table S4).

In addition to laboratory testing of a variety strains of Influenza A, Influenza B, RSV and SARS-CoV-2, *in silico* inclusivity analysis was performed, using GISAID, NCBI Virus, Geneious Prime and a Python script. For Flu A, it is predicted that at least 99.9% of sequences would be amplified and detected; for Flu B, 99.6%; and for RSV, 98.7%. For SC2, the analysis predicted that at least 99.99% of all available sequences would be amplified and detected by at least one of the three assays.

### Cross-reactivity

Cross-reactivity was evaluated by testing a panel of 40 microorganisms, comprising 3 human coronaviruses, 1 MERS coronavirus and 36 common pathogens or those potentially encountered in the nasopharynx (Table S5). None of the organisms tested had positive results for any viral target; the analytical specificity of Fast PCR MRP is 100%. In addition, *in silico* analysis (organism-specific BLAST) was performed and demonstrated that only the intended targets would be detected by the assay (no cross-reactivity).

### Interference

Interference was evaluated for 15 endogenous and exogenous substances in ANS matrix with a 3x LoD quadspike sample. None of the substances tested resulted in less than 100% positivity for all targets (Table S6). For the microbial interference evaluation, the same 44 organisms tested for cross-reactivity were evaluated for microbial interference using ANS matrix and a 3x LoD quadspike sample. None of the organisms caused microbial interference (defined as less than 100% positivity for each viral target; Table S5). Since the Fast PCR MRP is a multiplex test, we also evaluated competitive inhibition, which was not observed. The presence of one virus at a high concentration did not impact the ability of the test to detect the other viruses at 3x LoD.

### Quality Control

Instrument performance was monitored daily for each Fast PCR Operating Module by running the External Positive and External Negative Controls. Descriptive statistics for External Positive Control runs is shown in Table 5.

**Table 5:** External Positive Control Results.

| Target | N | Average Ct | %CV |
| --- | --- | --- | --- |
| Flu A | 127 | 35.1 | 3.5 |
| Flu B | 127 | 35.4 | 2.3 |
| RSV | 127 | 34.4 | 2.5 |
| SARS-CoV-2 | 127 | 34.8 | 1.8 |

## Discussion

Rapid and accurate detection and differentiation of common respiratory viruses is essential in infection control, prompt initiation of antiviral therapy, and ruling out other pathogens. This is best accomplished through use of RT-PCR based assays for detection of viral RNA as they are more sensitive than antigen detection assays. As an example, for one antigen test (Abbott BinaxNOW COVID-19 Ag card), the LoD was estimated to be between 4×10^4^ and 8×10^4^ copies/swab (equivalent to 8×10^5^ to 1.6×10^6^ copies/mL) (12), which is approximately 100-fold higher than several RT-PCR tests; subsequently, it is estimated that 46% of SARS-CoV-2 cases would have been missed if it were the only test available for screening (13). The AMDI Fast PCR MRP is a CLIA-waivable POC system that integrates sample preparation and RT-PCR for qualitative detection and differentiation of Flu A, Flu B, RSV and SARS-CoV-2 in ∼10 minutes.

The analytical sensitivity, demonstrated by the limit of detection of the assay, is a measure of the ability of the test to detect a pathogen in the case of low viral load or early-stage infection. Tests with high sensitivity (low LoD) are especially important for SARS-CoV-2 and other respiratory pathogens for control of outbreaks (14–15) as well as where low viral loads are possible, especially due to poor sample collection technique (16). The Fast PCR MRP test had strain-dependent LoDs in the range of 83 - 500 copies/mL, comparable to widely used point of care systems such as Cepheid Xpert Xpress/Flu/RSV+ or BIOFIRE SPOTFIRE Respiratory (R) Panel Mini. Further, the sensitivity established using analytical spikes of single viral targets was not affected when tested as a quadruple spiked specimen. This demonstrates the ability of the AMDI Fast PCR MRP to detect co-infections at very low titers of virus which is critical during respiratory viral season when more than one viral pathogen can be expected to be in circulation.

Reproducibility is an essential part of quality control and is necessary to determine if the test will perform as expected in different laboratories and environmental conditions with different operators. Testing for the Fast PCR MRP multi-site reproducibility study was performed at three CLIA-waived clinical sites. This study demonstrated reproducible results across a wide variety of users and instruments, as evidenced by the high levels of agreement for the samples and low variability in Ct value. Across all three sites, for each positive sample (2x LoD or 5x LoD), the %CV was less than 3.6%. In addition, the LoD study also demonstrated a high level of repeatability (CV < 4.0 %) at very low viral titers. Taken together, the Fast PCR MRP demonstrated excellent repeatability and reproducibility to be used as a reliable aid in overall patient management.

While the clinical agreement study (8) provides information about the test performance against circulating strains, it is essential to ensure that the test can detect clinically relevant subtypes and strains, especially with RNA viruses that are subject to evolution and mutation. Using a highly conserved target region is essential for the highest possible inclusivity. For this purpose, Fast PCR MRP targets the Matrix gene of the Flu A, Flu B and RSV genomes. For SARS-CoV-2, there are three targets (Nucleocapsid, ORF1AB and ORF8); detection of only one of these is required for a positive result. The use of at least two targets for SARS-CoV-2 was an early recommendation of the WHO (16). The inclusivity testing of the Fast PCR MRP confirmed that the assay design for each viral target in the multiplex test was robust to detect 100% of the available strains at 3X LoD and >98% of available genomic sequences *in silico* for each virus. Although this is not predictive of performance of the test with evolving strains, the inclusivity testing provides confidence that the Fast PCR MRP test is designed to prevent false negative results.

There are many respiratory viruses (other coronaviruses, parainfluenza viruses, rhinovirus, human metapneumovirus, adenoviruses, etc.) with similar symptoms, and it is important for a test to discriminate amongst these as well as Flu A, Flu B, RSV and SARS-CoV-2. Cross-reactivity was tested using quantified stocks of individual cultures as opposed to clinical samples that are positive for various organisms. The cross-reactivity evaluation was expanded beyond the organisms that were wet-tested by *in silico* analysis (organism-specific BLAST) of the oligonucleotides in the Fast PCR MRP kit. These studies demonstrated a lack of cross-reactivity to organisms other than the intended targets. Combined with the high NPA from the clinical study (8), this indicates a high level of specificity for Fast PCR MRP.

The Fast PCR MRP test was not found to have interference caused by any of the therapies and remedies, nor endogenous substances that were tested. The Fast PCR MRP test will produce reliable results even after relevant, common medications are used to manage respiratory symptoms. Neither blood nor mucus, common endogenous biological matrices in nasal swab samples, is expected to cause a false-negative result when using the Fast PCR MRP test. Interference was also not observed due to any of the organisms tested for microbial interference. In addition, the lack of competitive inhibition indicates that the Fast PCR MRP kit can identify samples with co-infections.

The analytical studies for the AMDI™ Fast PCR MRP demonstrate the appropriateness of this test for the sensitive and specific detection and differentiation of SARS-CoV-2, Flu A, Flu B and RSV. This test has advantages over many traditional NAATs and rapid antigen detection tests. The Fast PCR MRP test is highly sensitive and specific and provides results in approximately 10 minutes with a very simple workflow that does not require up-front processing, interpretation, or judgment on the part of the user. The simple and convenient nature of the Fast PCR MRP test will aid overall management of patients with symptoms of upper respiratory infections.

## Data Availability

All data produced in the present work are contained in the manuscript.

## Acknowledgements

We acknowledge the staff at the participating clinical sites, site coordinators at MDC Associates, Karen Copeland at Boulder Statistics LLC.

## Supplementary Tables

**Table S1:** LoD Results for Confirmation and Additional Levels.

| Target | Strain | Preliminary LoD (copies/mL) | Confirmatory LoD Results |  |  |  |  |
| --- | --- | --- | --- | --- | --- | --- | --- |
|  |  |  | Copies/mL | TCID<sub>50</sub>/mL | % Positivity | Average Ct | %CV |
| Flu A | A/Victoria/4897/2022 | 83 | 250 | 26.9 | 100% (20/20) | 38.5 | 2.4 |
|  |  |  | **83** | **9.0** | **100% (20/20)** | **40.8** | **3.7** |
|  |  |  | 42 | 4.5 | 65% (13/20) | 40.8 | 2.2 |
|  | A/Darwin/9/2021 | 250 | 750 | 2.4 | 100% (20/20) | 37.6 | 1.7 |
|  |  |  | **250** | **0.8** | **100% (20/20)** | **39.8** | **3.6** |
|  |  |  | 83 | 0.3 | 65% (13/20) | 40.9 | 3.9 |
| Flu B | B/Colorado/06/2017 | 83 | **500** | **2.4** | **100% (20/20)** | **39.2** | **1.5** |
|  |  |  | 250 | 1.2 | 85% (17/20) | 40.3 | 2.5 |
|  |  |  | 83 | 0.4 | 90% (18/20) | 41.3 | 2.3 |
|  | B/Phuket/3073/2013 | 250 | 750 | 4.6 | 100% (20/20) | 38.0 | 1.3 |
|  |  |  | **250** | **1.5** | **100% (20/20)** | **39.1** | **1.8** |
|  |  |  | 83 | 0.5 | 85% (17/20) | 40.9 | 1.9 |
| RSV | RSV A 2006 isolate | 250 | 750 | 0.8 | 100% (20/20) | 37.7 | 3.0 |
|  |  |  | **250** | **0.3** | **100% (20/20)** | **39.2** | **3.9** |
|  |  |  | 83 | 0.1 | 65% (13/20) | 41.2 | 3.4 |
|  | RSV B (Washington) | 250 | 750 | 1.1 | 100% (20/20) | 39.3 | 3.5 |
|  |  |  | **500** | **0.7** | **95% (19/20)** | **40.0** | **3.8** |
|  |  |  | 250 | 0.4 | 75% (15/20) | 40.3 | 1.9 |
| SC2 | USA/WA/1/2020 | 750 | 750 | 0.4 | 95% (19/20) | 37.6 | 1.3 |
|  |  |  | **500** | **0.2** | **100 % (20/20)** | **39.4** | **2.8** |
|  |  |  | 250 | 0.1 | 65% (13/20) | 39.6 | 2.9 |

**Table S2:**
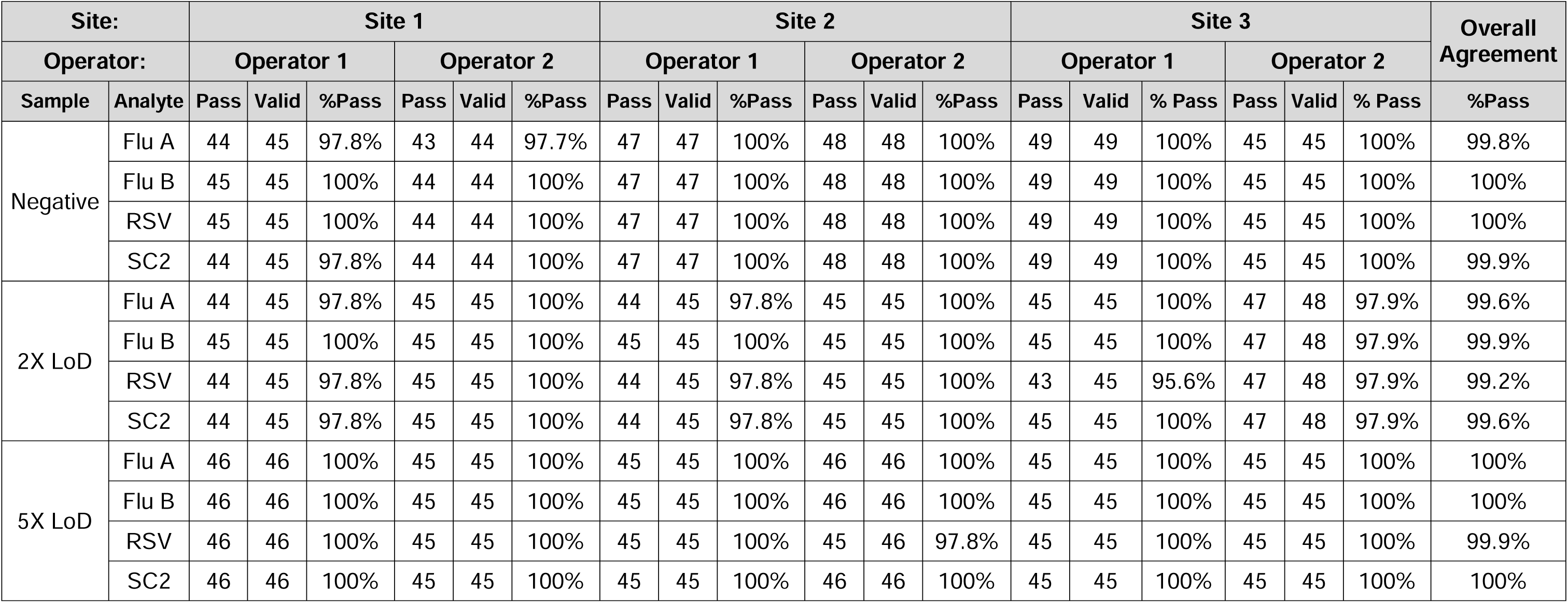
Fast PCR MRP Reproducibility Results Stratified by Testing Site.

| Site: |  | Site 1 |  |  |  |  |  | Site 2 |  |  |  |  |  | Site 3 |  |  |  |  |  | Overall Agreement |
| --- | --- | --- | --- | --- | --- | --- | --- | --- | --- | --- | --- | --- | --- | --- | --- | --- | --- | --- | --- | --- |
| Operator: |  | Operator 1 |  |  | Operator 2 |  |  | Operator 1 |  |  | Operator 2 |  |  | Operator 1 |  |  | Operator 2 |  |  | %Pass |
| Sample | Analyte | Pass | Valid | %Pass | Pass | Valid | %Pass | Pass | Valid | %Pass | Pass | Valid | %Pass | Pass | Valid | % Pass | Pass | Valid | % Pass |  |
| Negative | Flu A | 44 | 45 | 97.8% | 43 | 44 | 97.7% | 47 | 47 | 100% | 48 | 48 | 100% | 49 | 49 | 100% | 45 | 45 | 100% | 99.8% |
|  | Flu B | 45 | 45 | 100% | 44 | 44 | 100% | 47 | 47 | 100% | 48 | 48 | 100% | 49 | 49 | 100% | 45 | 45 | 100% | 100% |
|  | RSV | 45 | 45 | 100% | 44 | 44 | 100% | 47 | 47 | 100% | 48 | 48 | 100% | 49 | 49 | 100% | 45 | 45 | 100% | 100% |
|  | SC2 | 44 | 45 | 97.8% | 44 | 44 | 100% | 47 | 47 | 100% | 48 | 48 | 100% | 49 | 49 | 100% | 45 | 45 | 100% | 99.9% |
| 2X LoD | Flu A | 44 | 45 | 97.8% | 45 | 45 | 100% | 44 | 45 | 97.8% | 45 | 45 | 100% | 45 | 45 | 100% | 47 | 48 | 97.9% | 99.6% |
|  | Flu B | 45 | 45 | 100% | 45 | 45 | 100% | 45 | 45 | 100% | 45 | 45 | 100% | 45 | 45 | 100% | 47 | 48 | 97.9% | 99.9% |
|  | RSV | 44 | 45 | 97.8% | 45 | 45 | 100% | 44 | 45 | 97.8% | 45 | 45 | 100% | 43 | 45 | 95.6% | 47 | 48 | 97.9% | 99.2% |
|  | SC2 | 44 | 45 | 97.8% | 45 | 45 | 100% | 44 | 45 | 97.8% | 45 | 45 | 100% | 45 | 45 | 100% | 47 | 48 | 97.9% | 99.6% |
| 5X LoD | Flu A | 46 | 46 | 100% | 45 | 45 | 100% | 45 | 45 | 100% | 46 | 46 | 100% | 45 | 45 | 100% | 45 | 45 | 100% | 100% |
|  | Flu B | 46 | 46 | 100% | 45 | 45 | 100% | 45 | 45 | 100% | 46 | 46 | 100% | 45 | 45 | 100% | 45 | 45 | 100% | 100% |
|  | RSV | 46 | 46 | 100% | 45 | 45 | 100% | 45 | 45 | 100% | 45 | 46 | 97.8% | 45 | 45 | 100% | 45 | 45 | 100% | 99.9% |
|  | SC2 | 46 | 46 | 100% | 45 | 45 | 100% | 45 | 45 | 100% | 46 | 46 | 100% | 45 | 45 | 100% | 45 | 45 | 100% | 100% |

**Table S3:** Reproducibility Results for Fast PCR MRP Positive and Negative External Controls.

|  | Fast PCR MRP Positive Control |  |  |  |  |  |  |  | Fast PCR MRP Negative Control |  |  |  |  |  |  |  |
| --- | --- | --- | --- | --- | --- | --- | --- | --- | --- | --- | --- | --- | --- | --- | --- | --- |
| Fast PCR MRP Kit Lot | Lot 1 |  | Lot 2 |  | Lot 3 |  | All |  | Lot 1 |  | Lot 2 |  | Lot 3 |  | All |  |
|  | Pass | Fail | Pass | Fail | Pass | Fail | Total | %Pass | Pass | Fail | Pass | Fail | Pass | Fail | Total | %Pass |
| Site 1 | 8 | 0 | 7 | 0 | 9 | 0 | 24 | 100%<br>24/24 | 8 | 0 | 8 | 0 | 8 | 0 | 24 | 100%<br>24/24 |
| Site 2 | 6 | 0 | 5 | 0 | 6 | 0 | 17 | 100%<br>17/17 | 5 | 0 | 5 | 0 | 6 | 0 | 16 | 100%<br>16/16 |
| Site 3 | 6 | 0 | 6 | 0 | 6 | 0 | 18 | 100%<br>18/18 | 6 | 0 | 6 | 0 | 6 | 0 | 18 | 100%<br>18/18 |
| Total | 20 | 0 | 18 | 0 | 21 | 0 | 59 | 100%<br>59/59 | 19 | 0 | 19 | 0 | 20 | 0 | 58 | 100%<br>58/58 |
| %Pass | 100%<br>20/20 |  | 100%<br>18/18 |  | 100%<br>21/21 |  |  |  | 100%<br>19/19 |  | 100%<br>19/19 |  | 100%<br>20/20 |  |  |  |

**Table S4:**
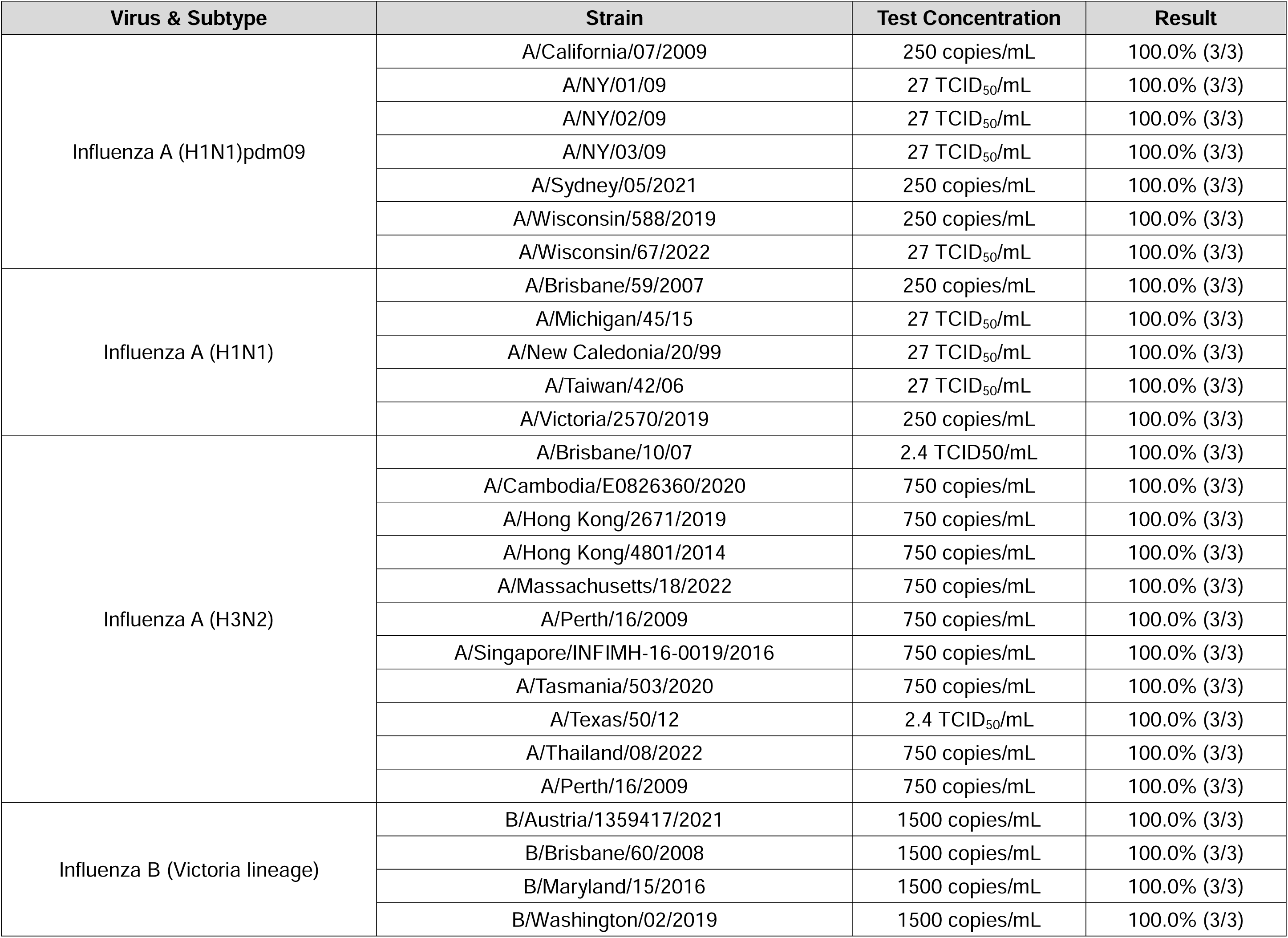
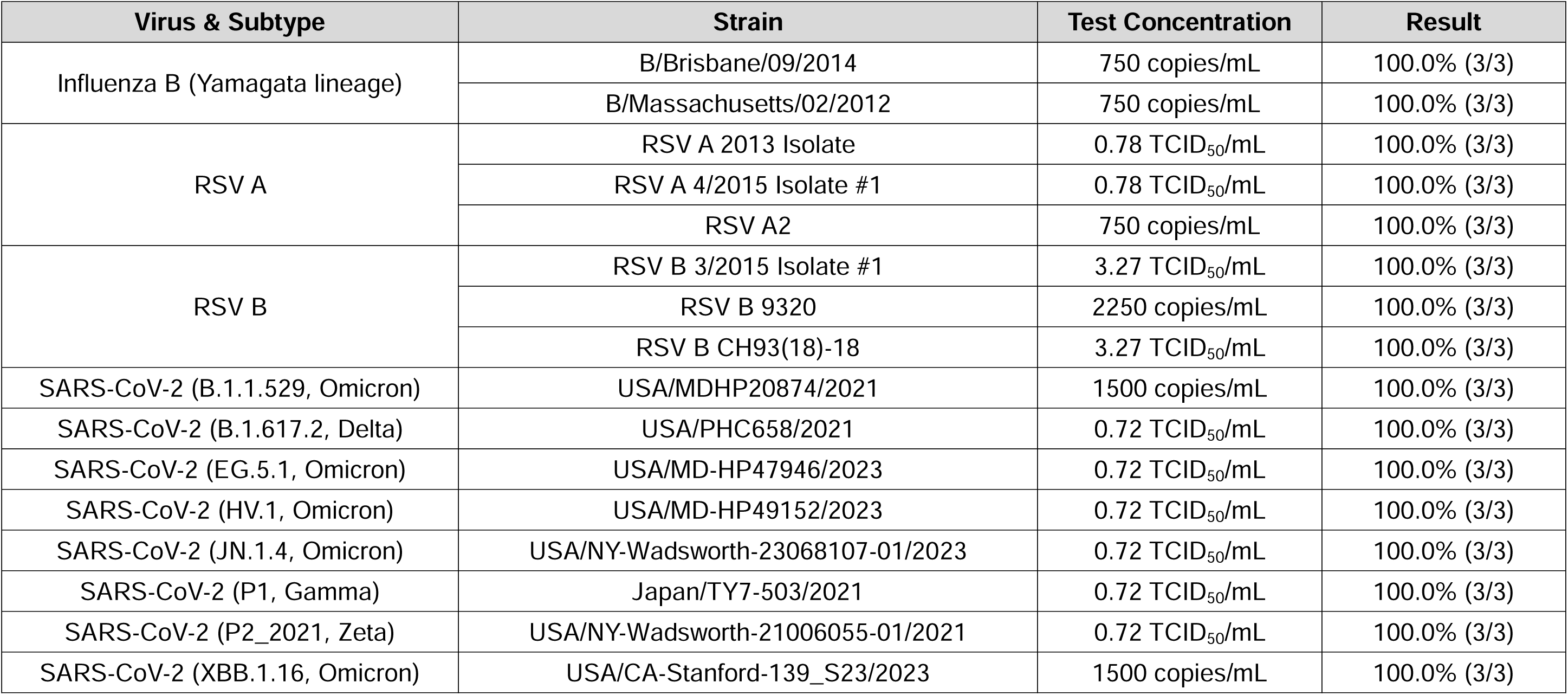
Results of Inclusivity Testing.

**Table S5:** Organisms for Cross-reactivity and Microbial Interference Testing.

| Viruses |  |  |  |
| --- | --- | --- | --- |
| Organism | Test Concentration | Organism | Test Concentration |
| Adenovirus Type 1 | 1x10<sup>5</sup> TCID<sub>50</sub>/mL | Measles virus | 1.68x10<sup>4</sup> TCID<sub>50</sub>/mL |
| Adenovirus Type 7 | 1x10<sup>5</sup> TCID<sub>50</sub>/mL | MERS-CoV | 1x10<sup>5</sup> genome equivalents/mL |
| Cytomegalovirus | 4.7x10<sup>3</sup> TCID<sub>50</sub>/mL | Mumps virus | 1x10<sup>5</sup> TCID<sub>50</sub>/mL |
| Enterovirus D68 | 1.67x10<sup>4</sup> TCID<sub>50</sub>/mL | Parainfluenza virus 1 | 1x10<sup>5</sup> TCID<sub>50</sub>/mL |
| Epstein-Barr virus | 1x10<sup>5</sup> copies/mL | Parainfluenza virus 2 | 1x10<sup>5</sup> TCID<sub>50</sub>/mL |
| Human coronavirus 229E | 1.39x10<sup>4</sup> TCID<sub>50</sub>/mL | Parainfluenza virus 3 | 1x10<sup>5</sup> TCID<sub>50</sub>/mL |
| Human coronavirus NL63 | 1.18x10<sup>4</sup> TCID<sub>50</sub>/mL | Parainfluenza virus 4 | 1x10<sup>5</sup> TCID<sub>50</sub>/mL |
| Human coronavirus OC43 | 1x10<sup>5</sup> TCID<sub>50</sub>/mL | Rhinovirus 1A | 1.39x10<sup>4</sup> TCID<sub>50</sub>/mL |
| Human Metapneumovirus (hMPV) | 6.27 x10<sup>3</sup> TCID<sub>50</sub>/mL | RSV A | 1x10<sup>5</sup> copies/mL |
| Influenza A/Victoria/4897/2022 | 1x10<sup>5</sup> copies/mL | RSV B | 1x10<sup>5</sup> copies/mL |
| Influenza B/Colorado 06/2017 | 1x10<sup>5</sup> copies/mL | SC2 USA/WA1/2020 | 1x10<sup>5</sup> copies/mL |
| Bacteria |  |  |  |
| Organism | Test Concentration | Organism | Test Concentration |
| *Bordetella parapertussis* | 1x10<sup>6</sup> CFU/mL | *Mycoplasma genitalium* | 1x10<sup>6</sup> CCU/mL |
| *Bordetella pertussis* | 1x10<sup>6</sup> CFU/mL | *Mycoplasma pneumoniae* | 1x10<sup>6</sup> CFU/mL |
| *Corynebacterium diphtheriae* | 1x10<sup>6</sup> CFU/mL | *Neisseria meningitidis* | 1x10<sup>6</sup> CFU/mL |
| *Escherichia coli* | 1x10<sup>6</sup> CFU/mL | *Neisseria elongata* | 1x10<sup>6</sup> CFU/mL |
| *Fusobacterium necrophorum* | 1x10<sup>6</sup> CFU/mL | *Pseudomonas aeruginosa* | 1x10<sup>6</sup> CFU/mL |
| *Haemophilus influenzae* | 1x10<sup>6</sup> CFU/mL | *Staphylococcus aureus* | 1x10<sup>6</sup> CFU/mL |
| *Lactobacillus plantarum* | 1x10<sup>6</sup> CFU/mL | *Staphylococcus epidermidis* | 1x10<sup>6</sup> CFU/mL |
| *Legionella pneumophila* | 1x10<sup>6</sup> CFU/mL | *Streptococcus pneumoniae* | 1x10<sup>6</sup> CFU/mL |
| *Moraxella catarrhalis* | 1x10<sup>6</sup> CFU/mL | *Streptococcus pyogenes* | 1x10<sup>6</sup> CFU/mL |
| *Mycobacterium tuberculosis* | 1x10<sup>6</sup> CFU/mL | *Streptococcus salivarius* | 1x10<sup>6</sup> CFU/mL |
| Fungi |  |  |  |
| Organism | Test Concentration | Organism | Test Concentration |
| *Aspergillus fumigatus* | 1x10<sup>6</sup> CFU/mL | *Candida albicans* | 1x10<sup>6</sup> CFU/mL |

**Table S6:** Interference results.

| Substance | Active Ingredients | Test concentration |
| --- | --- | --- |
| Antibacterial, systemic | Tobramycin | 4 µg/mL |
| Antibiotic, nasal ointment | Mupirocin | 10 mg/mL |
| Chloraseptic Lozenges | Menthol, Benzocaine | 1.5 mg/mL |
| Chloraseptic Sore Throat Spray | Phenol | 15% (v/v) |
| Fluticasone Propionate (Nasal Spray) | Fluticasone Propionate | 5% (v/v) |
| Homeopathic Nasal Wash (Alkalol) | Eucalyptol, menthol, thymol, camphor, benzoin; oils of wintergreen, spearmint, pine and cinnamon | 10% (v/v) |
| Human Whole Blood (EDTA collection tube) | N/A | 3% (v/v) |
| Mucin (bovine submaxillary gland, type I-S) | Purified mucin protein | 0.5% (w/v) |
| Nasal Drops | Phenylephrine | 15% (v/v) |
| Nasal Spray | Oxymetazoline | 12% (v/v) |
| Nasal Spray | Cromolyn | 15% (v/v) |
| Naso GEL (NeilMed) | Sodium hyaluronate, Aloe vera | 5% (v/v) |
| Tamiflu | Oseltamivir Phosphate | 5 mg/mL |
| Zanamivir | Zanamivir | 282 ng/mL |
| Zicam Cold Remedy Nasal Spray | *Galphimia glauca* 4x, *Luffa operculata* 4x, *Sabadilla* 4x | 5% (v/v) |

